# SIGMA METRICS ASSESSMENT AS QUALITY IMPROVEMENT METHODOLOGY IN A CLINICAL CHEMISTRY LABORATORY

**DOI:** 10.1101/2021.06.13.21258863

**Authors:** Monika Garg, Neera Sharma, Saswati Das

## Abstract

**Background:** The concept of sigma metrics & lean six sigma is well known in the field of healthcare. However not many labs utilize the six sigma metrics for maintenance of high quality laboratory performance. A minimum value of 3 σ is desired in any clinical laboratory & values of σ≥ 6 are regarded as gold standard for obtaining high quality lab reports.

**Aims &Objectives:** To calculate bias, cv & sigma metrics from the IQC & EQC data in order to ascertain extent of quality management in our lab.

**Materials &Methods:** An extensive study of sample processing and quality practices was carried out in the Central Laboratory of Department of Biochemistry; PGIMER &Dr. RML Hospital, New Delhi; from Feb 2020 to July 2020. The IQC used(both level I & level II) were from Biorad Laboratories India (lyphochek assayed chemistry control) & the EQC used was from Randox Laboratories, UK. All the controls were run on Beckman Coulter clinical chemistry analyser AU 680. Total 14 clinical parameters were analysed & subsequently; Mean, S.D., CV, bias & σ were calculated through their respective formulas.

**Results:** Sigma level was more than 6 for both levels of IQC was observed for Amylase. It indicates world class performance. Total bilirubin, AST, Triglyceride & HDL depicted σ values between 3.1 – 6 for both L_1_ & L_2._ Iron showed σ value of 5.5 in L_1_ whereas it was 3.78 in L_2_.

**Conclusion:** Sigma metrics in clinical laboratory is an essential technique to ascertain poor assay performance, along with assessment of the efficiency of existing laboratory process.

## Introduction

In any Healthcare Laboratory, the term “quality” is defined as conformance to the requirements of users (nurses & physicians) or customers (patients or other parties who pay the bills) & the satisfaction of their needs & expectations.^1^

Certain good quality indicators like reduced number of repeats & reruns; reduced time for sample transportation and storage ultimately leading to decreased Turn Around Time (TAT) etc. signify the good quality of Lab reports.^2,3^ A good quality laboratory performance is depicted in the test reports it generates and is also equally reflected in the Quality Controls performed as its performance checks. Besides Internal Quality Control (IQC) and External Quality Control (EQC); their exhaustive interpretation has become quite indispensable in the present scenario. This can be carried out by using the concept of “Six Sigma” and “Lean Six Sigma”. In spite of being introduced in Motorola since 1986; “Six Sigma” methodology was adopted in laboratory medicine in the year 2000.^4^This concept basically revolves around decreasing the “non-value-adding” steps of a process and to enhance the quality of the process by reducing errors associated with it.^5^

Sigma (σ) reflects the Defects or errors per Million Opportunities (DPMO). The sigma refers to the number of SDs from the mean a process can be, before it is outside the acceptable limits. The process having 6σ is considered extremely precise, having only 3.4 DPMO.

## Materials &Methods

An exhaustive study of sample processing and quality control procedures was carried out in the Central Laboratory of Department of Biochemistry; PGIMER &Dr. RML Hospital, New Delhi; from Feb 2020 to July 2020. The corresponding data was collected and analysed subsequently.

The IQC used (both level I & level II) were from Biorad Laboratories India (lyphochek assayed chemistry control) & the EQC used was from Randox Laboratories, UK. All the controls were run on Beckman Coulter clinical chemistry analyser AU 680. On each day, both the levels of IQC were run & analysed. The patients’ samples were run & reported only if the IQC came within the acceptable range following the Westgard rules.

The parameters which were analysed include Glucose, Urea, Total bilirubin, AST, ALT, total protein, albumin, cholesterol, triglyceride, High Density Lipoprotein (HDL), sodium, potassium, amylase & Iron.

By using IQC data; Mean & S.D. were determined. Subsequently, CV% (coefficient of variation) was calculated using the following formula:-

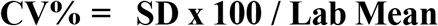

Further; Bias was ascertained by employing RIQAS with the following formula:-

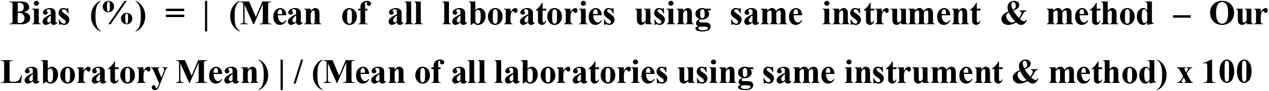

**Values for TE_a_ (Total allowable Error)** were taken from **CLIA** (Clinical Laboratories Improvement Act) guidelines

Finally, Sigma metrics (σ) was determined using following equation:-

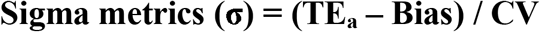

The methodology employed to implement Lean Six Sigma in our Central Laboratory was DMAIC i.e. Define, Measure, Analyse, Improve and Control. It means “defining” the problem due to which our results are deviating from the range of established standards. It also includes defining the resources which may be required to solve the problem.^6^

Subsequently, in order to solve the problem;its extent was “measured”, through collection of pertinent data & retain it in a more presentable form e.g.collection of IQC & EQC data & calculating CV% & Bias% respectively from it.^6,7^

Further, the data was “analysed” in order to ascertain the root causes of the defect or problem.In this phase; the differences between our results & the target values are estimated along with determination of their possible causes.^8^

After that, the root causes were eliminated by implementing certain corrective measures in accordance with Westgard sigma rules^9^ to “improve” the process performance. Once the defect is rectified; certain preventive measures were also taken in order to “control” or check that such problem or defect should not occur in future.

In order to eliminate any errors and wasteful steps during sample processing; every step of sample processing was carefully reviewed and monitored. Many corrective measures were taken to reduce errors at pre-analytical; analytical and post-analytical phases in order to reduce TAT and improve quality of sample processing.

## Results &Observations

In the present study IQC & EQC data of 6 months (February 2020 to July 2020) was collected & compiled in excel sheets to calculate Mean, S.D., CV%, Bias and Sigma Metrics for 14 different parameters. While evaluating IQC,it can be observed that only Total bilirubin, ALT & HDL had CV% >5 in L_1_ and rest all parameters showed CV%<5(Table 1). As far as L_2_ is concerned, except for HDL (CV%>5); rest all parameters had CV%<5. Now taking EQC into consideration, out of all the measured parameters, average bias of sodium was minimum (0.85) whereas Triglyceride & HDL had bias >5. (Table 2)

**Table 1.** CV% calculated from Internal Quality Control L_1_ & L_2_from February 2020 to July 2020 (6 months).

|  | L<sub>1</sub> |  |  |  |  |  |  | L<sub>2</sub> |  |  |  |  |  |  |
| --- | --- | --- | --- | --- | --- | --- | --- | --- | --- | --- | --- | --- | --- | --- |
| Parameter | Feb | March | April | May | June | July | Average | Feb | March | April | May | June | July | Average |
| Glucose | 2.43 | 1.78 | 2.18 | 3.76 | 4.81 | 4.29 | 3.21 | 2.46 | 1.63 | 2.68 | 4.54 | 5.51 | 3.78 | 3.43 |
| Urea | 2.86 | 4.22 | 3.77 | 1.52 | 3.26 | 4.32 | 3.32 | 3.08 | 2.77 | 3.5 | 2.64 | 3.35 | 4.8 | 3.36 |
| Total Bilirubin | 4.83 | 3.88 | 5.36 | 2.04 | 5.63 | 9.03 | 5.13 | 2.22 | 4.44 | 4.54 | 2.94 | 4.09 | 6.79 | 4.17 |
| AST | 3.28 | 4.16 | 3.88 | 2.62 | 4.27 | 4.75 | 3.83 | 2.14 | 2.01 | 3.12 | 2.87 | 3.48 | 4.39 | 3.0 |
| ALT | 8.09 | 9.19 | 7.12 | 3.75 | 5.06 | 5.24 | 6.41 | 2.94 | 2.93 | 5.91 | 3.09 | 3.26 | 4.64 | 3.8 |
| Total Protein | 3.12 | 2.27 | 3.15 | 4.01 | 3.79 | 3.35 | 3.28 | 3.39 | 2.55 | 4.21 | 5.17 | 4.05 | 3.43 | 3.8 |
| Albumin | 2.16 | 1.96 | 2.27 | 1.7 | 3.85 | 5.59 | 2.92 | 2.55 | 1.74 | 2.89 | 3 | 3.36 | 5.49 | 3.17 |
| Cholesterol | 2.43 | 4.36 | 3.29 | 2.31 | 2.31 | 3.59 | 3.05 | 2.03 | 2.42 | 2.86 | 2.6 | 2.14 | 4.67 | 2.79 |
| Triglyceride | 6 | 2.35 | 4.14 | 2.66 | 4.06 | 5.03 | 4.04 | 5.52 | 2.68 | 3.04 | 3.67 | 5.8 | 6.21 | 4.49 |
| HDL | 4.48 | 5.98 | 6.42 | 5.25 | 6.49 | 5.14 | 5.63 | 3.53 | 4.09 | 5.67 | 6.14 | 6.78 | 6.25 | 5.41 |
| Sodium | 1.34 | 1.19 | 1.72 | 1.31 | 1.31 | 2.19 | 1.51 | 1.16 | 1.31 | 2.52 | 1.56 | 1.18 | 2.25 | 1.66 |
| Potassium | 1.53 | 1.71 | 2.08 | 1.87 | 1.53 | 2.46 | 1.86 | 1.28 | 1.33 | 2.77 | 1.51 | 1.58 | 2.54 | 1.84 |
| Amylase | 2.22 | 3.15 | 2.92 | 2.45 | 2.01 | 3.47 | 2.7 | 1.91 | 1.34 | 4.29 | 2.98 | 2.58 | 4.48 | 2.93 |
| Iron | 2.86 | 3.59 | 3.75 | 2.63 | 2.24 | 3.44 | 3.08 | 3.63 | 4.73 | 4.29 | 5.6 | 3.88 | 4.76 | 4.48 |

**Table 2.** Bias % calculated from RIQAS for 6 months (February 2020 to July 2020).

| Parameter | Feb | March | April | May | June | July | Average |
| --- | --- | --- | --- | --- | --- | --- | --- |
| Glucose | 1.36 | 3.98 | 1.32 | 6.17 | 3.14 | 3.61 | 3.26 |
| Urea | 0.07 | 0.9 | 4.9 | 0.68 | 3.65 | 3.15 | 2.22 |
| Total Bilirubin | 0 | 1.79 | 5.67 | 0 | 1.79 | 0.59 | 1.64 |
| AST | 2.78 | 0.76 | 3.3 | 2.44 | 3.57 | 10.08 | 3.82 |
| ALT | 0.83 | 2.9 | 2.2 | 9.09 | 2.24 | 6 | 3.87 |
| Total Protein | 2.63 | 1.72 | 9.09 | 3.45 | 0 | 0.87 | 2.96 |
| Albumin | 0.48 | 2.6 | 3.7 | 0 | 1.96 | 2.25 | 1.83 |
| Cholesterol | 5.24 | 6.01 | 2.75 | 2.09 | 4.56 | 3.49 | 4.02 |
| Triglyceride | 7.8 | 0.39 | 3.2 | 6.5 | 10.04 | 3.59 | 5.25 |
| HDL | 6.2 | 3.84 | 3.92 | 0 | 13.50 | 3.39 | 5.14 |
| Sodium | 1.29 | 0.97 | 0.06 | 1.05 | 0.8 | 0.98 | 0.85 |
| Potassium | 2.44 | 2.24 | 0.17 | 2.56 | 1.59 | 0.51 | 1.58 |
| Amylase | 5.06 | 1.28 | 0.64 | 0.61 | 1.54 | 0.12 | 1.54 |
| Iron | 4.09 | 4.04 | 3.01 | 2.74 | 2.09 | 2.48 | 3.07 |

As seen in (Table3)average σ value for amylase is maximum for both levels of IQC.

According to (Table 4); σ>6 for both levels of IQC was observed for Amylase. It indicates world class performance of this analyte. Further, 4 parameters namely Total bilirubin, AST, Triglyceride & HDL depicted σ values between 3.1 – 6 for both L_1_ & L_2._ Iron showed σ value of 5.5 in L_1_ but 3.78 in L_2_. Remaining all parameters had σ<3 in L_1_. As far as L_2_ is concerned, besides ALT which had σ value 4.24; rest all analytes had σ<3.

**Table 3.** Sigma metrics calculated from Internal Quality Control L_1_ &L_2_from February 2020 to July 2020 (6 months).

| Parameter | L<sub>1</sub> |  |  |  |  |  |  | L<sub>2</sub> |  |  |  |  |  |  |
| --- | --- | --- | --- | --- | --- | --- | --- | --- | --- | --- | --- | --- | --- | --- |
|  | Feb | March | April | May | June | July | Average | Feb | March | April | May | June | July | Average |
| Glucose | 3.54 | 3.4 | 3.9 | 1 | 1.42 | 1.5 | 2.46 | 3.5 | 3.7 | 3.23 | 3.5 | 1.2 | 1.7 | 2.8 |
| Urea | 3.11 | 1.9 | 1.1 | 5.4 | 1.64 | 1.3 | 2.4 | 2.9 | 2.9 | 1.17 | 3.1 | 1.6 | 1.2 | 2.1 |
| Total Bilirubin | 4.3 | 4.7 | 2.7 | 9.7 | 3.2 | 2.1 | 4.45 | 8.9 | 4.1 | 3.1 | 6.7 | 5.3 | 3 | 5.18 |
| AST | 5.2 | 4.6 | 4.3 | 6.7 | 3.8 | 2.1 | 4.45 | 8.01 | 9.5 | 5.3 | 6.1 | 4.7 | 2.3 | 5.98 |
| ALT | 2.3 | 1.8 | 2.5 | 2.9 | 3.5 | 2.7 | 2.61 | 6.5 | 5.8 | 3 | 3.5 | 5.4 | 3 | 4.53 |
| Total Protein | 2.35 | 3.6 | 0.3 | 1.6 | 2.6 | 2.7 | 2.19 | 2.1 | 3.2 | 0.2 | 1.3 | 2.4 | 2.6 | 1.96 |
| Albumin | 4.4 | 3.8 | 2.8 | 5.8 | 2.1 | 1.4 | 3.38 | 3.7 | 4.3 | 2.2 | 3.3 | 3.6 | 1.4 | 3.08 |
| Cholesterol | 1.9 | 0.9 | 2.2 | 3.4 | 2.3 | 1.8 | 2.08 | 2.3 | 1.6 | 2.5 | 3 | 6.8 | 1.4 | 2.93 |
| Triglyceride | 2.8 | 10.4 | 5.3 | 6.9 | 3.7 | 4.2 | 5.55 | 3.1 | 9.1 | 7.1 | 8.6 | 6 | 4.6 | 6.41 |
| HDL | 5.2 | 4.4 | 4 | 5.7 | 2.5 | 5.2 | 4.5 | 6.7 | 6.4 | 4.6 | 4.9 | 2.4 | 4.2 | 4.86 |
| Sodium | 2.7 | 3.3 | 2.8 | 3 | 3.2 | 1.8 | 2.8 | 3.2 | 3.1 | 1.9 | 3.8 | 4.9 | 1.8 | 3.11 |
| Potassium | 2.3 | 2.2 | 2.8 | 1.8 | 2.8 | 2.2 | 2.35 | 2.8 | 2.8 | 2.1 | 5.6 | 4.8 | 2.1 | 3.36 |
| Amylase | 11.2 | 9.1 | 10 | 11.9 | 14.1 | 8.6 | 10.81 | 13.1 | 21.4 | 6.8 | 9.8 | 11 | 6.7 | 11.46 |
| Iron | 5.5 | 4.4 | 4.5 | 6.5 | 7.9 | 5.1 | 5.65 | 4.4 | 3.4 | 3.9 | 4 | 5.7 | 3.7 | 4.18 |

**Table 4.** Average Bias, Average CV% & sigma metrics calculated for 6 months for both levels of IQC (L_1_ & L_2_).

| Parameter | TE<sub>a</sub> % | Average Bias% | Average CV% (L<sub>1</sub>) | σ (L<sub>1</sub>) | Average CV% (L<sub>2</sub>) | σ (L<sub>2</sub>) |
| --- | --- | --- | --- | --- | --- | --- |
| Glucose | 10 | 3.26 | 3.21 | 2.09 | 3.43 | 1.96 |
| Urea | 9 | 2.22 | 3.32 | 2.04 | 3.36 | 2.01 |
| Total Bilirubin | 20 | 1.64 | 5.13 | 3.58 | 4.17 | 4.4 |
| AST | 20 | 3.82 | 3.83 | 4.22 | 3.0 | 5.39 |
| ALT | 20 | 3.87 | 6.41 | 2.51 | 3.8 | 4.24 |
| Total Protein | 10 | 2.96 | 3.28 | 2.14 | 3.8 | 1.85 |
| Albumin | 10 | 1.83 | 2.92 | 2.79 | 3.17 | 2.58 |
| Cholesterol | 10 | 4.02 | 3.05 | 1.95 | 2.79 | 2.14 |
| Triglyceride | 25 | 5.25 | 4.04 | 4.88 | 4.49 | 4.39 |
| HDL | 30 | 5.14 | 5.63 | 4.41 | 5.41 | 4.59 |
| Sodium | 5 | 0.85 | 1.51 | 2.74 | 1.66 | 2.5 |
| Potassium | 6 | 1.58 | 1.86 | 2.36 | 1.84 | 2.4 |
| Amylase | 30 | 1.54 | 2.7 | 10.5 | 2.93 | 9.71 |
| Iron | 20 | 3.07 | 3.08 | 5.5 | 4.48 | 3.78 |

## Discussion

In the process of maintaining high laboratory quality standards, six sigma is regarded as an indispensable tool. The concept of Lean six sigma aims at reducing the wasteful activities during sample processing. When sigma metrics is ≥ 6, then the process is said to have only 3.4 DPMO & it is regarded as the “World Class Quality”.

Although achievement of sigma metrics value 6 or more is not easy, but with appropriate precautions to minimise the errors associated with sample processing (at pre-analytical, analytical & post analytical phases); this goal can be approached.

In the present study, IQC & EQC data of 6 months (February 2020 to July 2020) was used to calculate Mean, S.D., CV%, bias & sigma metrics of 14 analytes. The two entities, S.D. & CV% are used to measure the extent of deviation & variation respectively of IQC test result with respect to its mean. In general, CV% is the favoured mode of presentation. If CV is less than 5%, then the particular method used for determination of an analyte’s concentration is said to have a very good performance& precise.^7,10,13^It can be visualised from Table1 & Table 2 that except for HDL, (CV%>5 in both L_1_& L_2_) along with Total bilirubin & ALT (having CV%>5 in L_1_); rest all parameters depicted CV<5%. This clearly indicates that our lab has achieved high level of precision in remaining 11 analytes.

Another important calculated index in the present study is bias% by using the EQC data. As per the definition, the term “bias” implies any discrepancy between the results of our lab & the peer group labs employing the same instrument & methodology.^7,11^ This means that we should try to minimise the bias, in order to decrease the differences between the lab results. Out of all the parameters measured in the present study, Sodium had the minimum bias of 0.85 (Table 2) while rest analytes had bias<5% (except HDL & Triglyceride). This shows high degree of accuracy in our lab results.

TE_a_ is the sum of random error (imprecision) and systematic error (bias or inaccuracy).^7,12,13^ Also, this term encompasses the pre□analytical variations, biologic variations etc that leads to variability in patients’ results.

In the present study; σ>6 for both levels of IQC was observed for Amylase (Table 4). Hence it required only 1_3s_ Westgard sigma rule to be followed since it was showing excellent performance. Among others, Total bilirubin, AST, Triglyceride & HDL depicted σ values between 3.1 – 6 for both L_1_ & L_2_with Iron showing σ value of 5.5 in L_1_and 3.78 in L_2_. It implies Westgardsigma multirole application is needed for such parameters. For parameters having σ values between 5 to 6; three rules namely 1_3s_,2_2s_, and R_4s_are needed.4-sigma quality requires addition of a 4^th^ rule and implementation of a 1_3s_/2_2s_/R_4s_/4_1s_ multirole. Those parameters having σ<3 requires extensive evaluation in terms of reducing analytical bias & imprecision.^9,11.12,14^

## Conclusion

Sigma metrics in clinical laboratory is a vital methodology to identify & correct any deviation of lab results from the prescribed standards. It can help us ascertain poor assay performance along with assessment of the efficiency of existing laboratory process. The unnecessary & time consuming wasteful additional steps can be eliminated using the concept of sigma metrics & lean six sigma. This will decrease the TAT & help in dispatching good quality reports for better patient management. Further, sigma metrics can help in devising appropriate strategies for the judicious utilisation of IQC & EQC in a large sized clinical laboratory.

## Data Availability

The tables containing the monthly IQC and EQC comparisons is added.

